# American Covid: An Econometric Analysis of Variants

**DOI:** 10.1101/2022.06.09.22276226

**Authors:** James McIntosh

**Affiliations:** Economics Department, Concordia University, 1455 de Maisonneuve Blvd. W. Montreal H3G 1M8, Quebec, Canada

**Keywords:** Covid-19, United States, Variants, Immunity, Econometric models

## Abstract

Covid-19 time series data on new cases from the United States for the period January 20, 2020 to February 7, 2022 is analyzed using a distributed lag econometric model applied to the Wild Type, Alpha, and Delta variants to determine the relative efficacy of vaccines and previous infection induced immunity. The results from this study confirm, for the most part, what others have found using large cross section samples: vaccines are effective in dealing with all three variants, more so with Delta. Infection induced immunity is always greater than or equal that delivered by a vaccination and is largest for the Delta variant. However, in the Delta case previous non-Delta infections had no prophylactic value in preventing new Delta infections.

## Introduction

The Covid 19 (SARS-covid-2) pandemic reached its second anniversary in March 2022. Health practitioners and medical experts have learned much about treating this virus and the pharmaceutical industry has produced a group of very effective vaccines in an amazingly short time. On the other, hand there is much that remains to be discovered about the properties of the virus. While results on initial vaccine efficacy are fairly clear for the Wild type variant but less so for other variants and there is much uncertainty about the duration of the effectiveness of these vaccines and how immunity induced by vaccines compares with immunity due to a previous infection. Moreover, for infection induced immunity it is not clear how long this lasts or how this varies across the three variants considered here.

Unobservable characteristics also play an important role in the analysis of Covid 19. When the virus was first observed in early 2000 it was thought that asymptomatic case were not important. This turned out not to be true^1^ and lead to policies which failed to accurately asses the risks of contagion by not using masks or social distancing. As new variants emerged the ability of confirmed new cases to accurately predict the actual number of new cases declined and now with the omicron variant researchers have very little idea of how many cases there actually are. Although reported new cases do not represent all cases they deserve to be analyzed because those who get tested and report their status are the most affected by the virus and, as such, deserve special consideration. As demonstrated below this data can also be used in models which allow for the effect of unobservables to be considered.

In addition to these challenging problems, as the data on new cases displayed in Figure 1 shows, there are a number of additional complexities that need to be examined. First, the data is highly non-linear with cycles with increasing amplitudes as well as positive trend. Because both the cumulative number of cases as well as the share of the population vaccinated are monotonic time series they will not be able to explain new cases unless there are other variables which can explain the cyclical components in the data. In the regression model that is used below to explain new cases a polynomial function of time is used to account for the cyclical nature of the data. The high frequency variation in the data also needs to be explained. This is captured by dummy variables representing each day.

**Figure 1.**
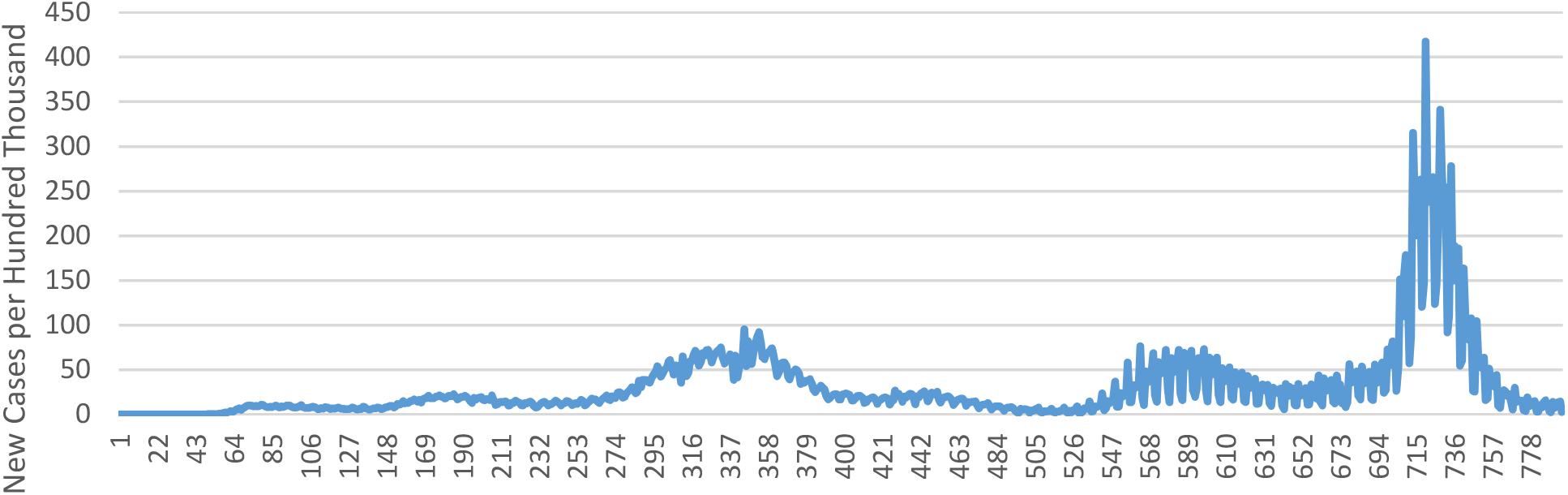
New Cases Per Hundred Thousand, January 22, 2000 to March 26, 2022 USA, All Variants

In the data of Figure 1 all the variants which contribute to the aggregate have a finite duration with a starting date and an end date. The Wild Type started on January 20, 2000 and ended on August 23, 2021. But there are important sub-intervals which are generated by the arrival of new variants. The alpha variant started in the United States on December 22, 2020 and along with the Delta variant which arrived on April 19, 2021 contributed to the demise of the Wild type variant. Thus the analysis of individual variants needs to take into account of the presence of other variants because the variant which is most infectious, as the literature on multiple virus SIR models shows, eliminates the other variants. If this is not done reductions in new case numbers could erroneously be attributed to the efficacy of vaccines. In addition, statistical problems arise in the estimation of the regression parameters when the variable associated with them is not present in all sub-periods. These issues are discussed in the last section.

The paper has the following format. In the next section contains a brief summary of what other researches have found. Section 3 describes the statistical model that is used to analyze the data. Section 4 contains the results and the paper ends with a discussion of them.

### A brief Review of the Literature

All of the literature discussed here is based on the analysis of large samples of randomly chosen individuals which are compared at two points in time to see how efficacy varies over sub groups like vaccinated individuals vs. individuals with a positive antibody test. Initially, vaccines were very effective. Clinical trials carried out in the latter half of 2000 on respondents exposed to the Wild type variant showed efficacy rates for both Moderna and Pfizer-BioNTech at around 95%, Pfizer (2012), Baden, Lindsey R, Hana M. El Sahly, Brandon Essink et al (2022), and Polack F.P., Thomas S.J., Kitchin N. et al (2020). Rates for Jonson & Johnson were lower at 66.9%, Sadoff, J, Glenda Gray, An Vandenbusch et al (2022). Similar results for New York State appear in Rosenberg, Eli S, Vajeera Dorabawila, Delia Easton et al (2022). As new variants emerged efficacy rates declined. For the Alpha variant the efficacy rates for Pfizer-BioNTech and Moderna were 76% and 81%, respectively in July 2001. Puranik, Arjun, Patrick J. Lenehan1, Eli Silvert et al (2021). For the Delta variant Patel, R., Mohamad Kakia, S. Venkat et al (2022) quote papers which show rates for Israel at 40.5%, the US at 75.0%, and the UK at 88.0%.

While efficacy is important so is the duration of immunity that previous infections generate. There is not much information on this issue but one study, Radbruch and Chang (2021) suggested that immunity due to previous infection is quite long lasting. Abu-Raddad, Chemaitelly and Coyle (2021) showed similar results for the Pfizer-BioNTech with a 95% efficacy rating for at least seven months for Qatar. Chemaitelly, Hassan, and Tang (2021) were not so optimistic about the immunological benefits of previous infections showing that this declined to 20% after seven months.

Some research has been devoted to the comparison of duration immunity generated by vaccination as opposed to previous infection. Another study by the Qatar team, Chemaitelly, Bertolini and Abu-Raddad (2021), showed that the immunological benefits from natural infection are slightly but significantly larger than those provided by vaccination for both the Alpha and Beta variants. More positive results were found in the survey by Kojima, N., N. K. Shrestha, & J. D. Klausner (2021) with countries with large sample sizes. Members of the Qatar research team also examined the protective effects of previous infection by variant. They showed while the while the Alpha, Beta and Delta variants had high own efficacy rates the Omicron variant was just over 50% effective. However, the efficacy of these variants with respect to other variants, like how well does having had an Alpha infection protect against getting a Delta infection, is unknown and is the main focus of attention of this research.

Many studies show high efficacy for vaccines against the wild Type and Alpha but efficacy declines significantly against delta and Omicron, Patel, R., Mohamad Kakia, S. Venkat et al (2022).

### An Econometric Model of Observed New Covid Cases

The statistical procedures employed here are used to investigate the relationship between newly observed cases of Covid-19 and the accumulated proportion of individuals vaccinated, the total number of recorded cases by variant, and a policy intervention variable, the Oxford Stringency Index, x_t_, and as well as cubic polynomial function, p_t_. This is included to account for exogenous trends and non-linear aspects of the data. Daily dummy variables are also included to take into account the consequences of recording errors.

The main variables in the model are n_vt_, the number of observed new cases and T_vt_ for the cumulative number of observed cases for variant v where v = w, α, δ. V is the cumulative percentage of the population with at least two vaccinations.

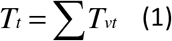

is the total cumulative number of observed cases. If n_t_ is the total number of new cases then

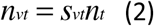

where s_vt_ is the share of variant v in new cases and

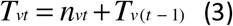

determines the evolution of the total number of cases for variant v.

The regression equation that is used to determine the relation between new cases and the other variables that are available as regressors is the geometric distributed lag model, Greene (2008, p. 677), of the form

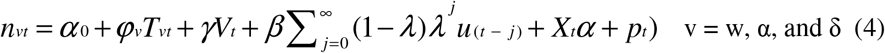

This is similar to the model used in McIntosh (2021) except for presence of the polynomial function in equation (4) and linear functions instead of the logarithms of the unlagged vaccination rates. The idea here is that new cases are generated by contact with infected individuals prior to their testing positive for Covid-19. The underlying assumption is that the earlier the individual is infected the lower the risk that this person infects others; u_(t-j)_ is the number of individuals that were infected at time (t-j) and 0<λ<1 is a decay parameter.

In equation (4) the stocks of cases, T_vt_, do not include unreported cases and unreported cases affect the number of new cases so a correction has to be made in estimation procedure. This is a measurement error problem and an instrumental variable (IV) procedure can be used to account for the unobservable component in the stock of cases. See Greene (2007, Ch. 12) for the details. The procedure regresses T_vt_ on a set of instruments (exogenous variables) and uses the fitted values of T_vt_ as regressors in equation (4). The data comes from Our World in Data (2022) which provides time series on n_t_, T_vt_, s_vt_, and x_t._

The analysis is carried out on each variant. This means that a particular time period is used for each variant in the estimation of the parameters in equation (4). The time periods are defined by the dates in Table 1.

**Table 1.**
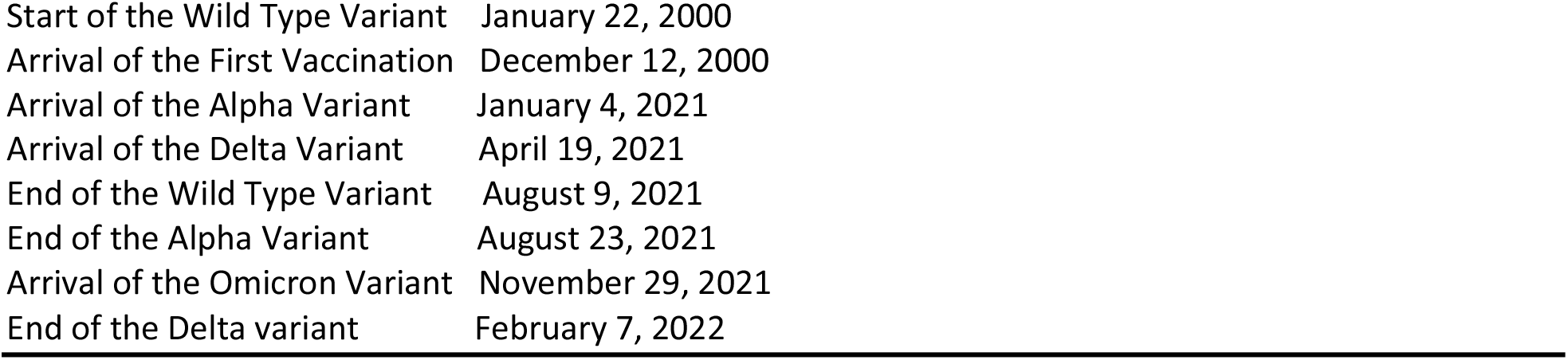
Important Dates in Covid 19 Pandemic.

|  |  |
| --- | --- |
| Start of the Wild Type Variant | January 22, 2000 |
| Arrival of the First Vaccination | December 12, 2000 |
| Arrival of the Alpha Variant | January 4, 2021 |
| Arrival of the Delta Variant | April 19, 2021 |
| End of the Wild Type Variant | August 9, 2021 |
| End of the Alpha Variant | August 23, 2021 |
| Arrival of the Omicron Variant | November 29, 2021 |
| End of the Delta variant | February 7, 2022 |

In addition, there are important sub-periods that need to be defined with respect to each variant. So, for example, in the analysis of new Wild Type cases three sub-periods have to be considered because all of the possible explanatory variables were not present for the full duration of the variant; vaccines started in December and both Alpha and Delta arrived at separate times before the end Wild Type.

## Results

The results from this study confirm, for the most part, what others have found: vaccines are effective in dealing with all three variants, more so with Delta. Infection induced immunity is always greater than or equal that delivered by a vaccination and is largest for the Delta variant. However, it should be noted that the estimate of the parameter associated with T_vt_ is often not significant.

The estimates of the parameters of interest for the model described by equation (4) appear in Table 2. There are results for the three American variants. Starting with the results on the Wild Type variety, vaccines became available only at the beginning of the second sub-period and had significant regression coefficients for vaccines which were the same for sub-periods two and three. Previous cases of the Wild type had no significant impact on new cases in any sub-period although they are similar in size to the vaccination parameters. This is possibly due to the inefficiency of the IV estimator.

**Table 2.**
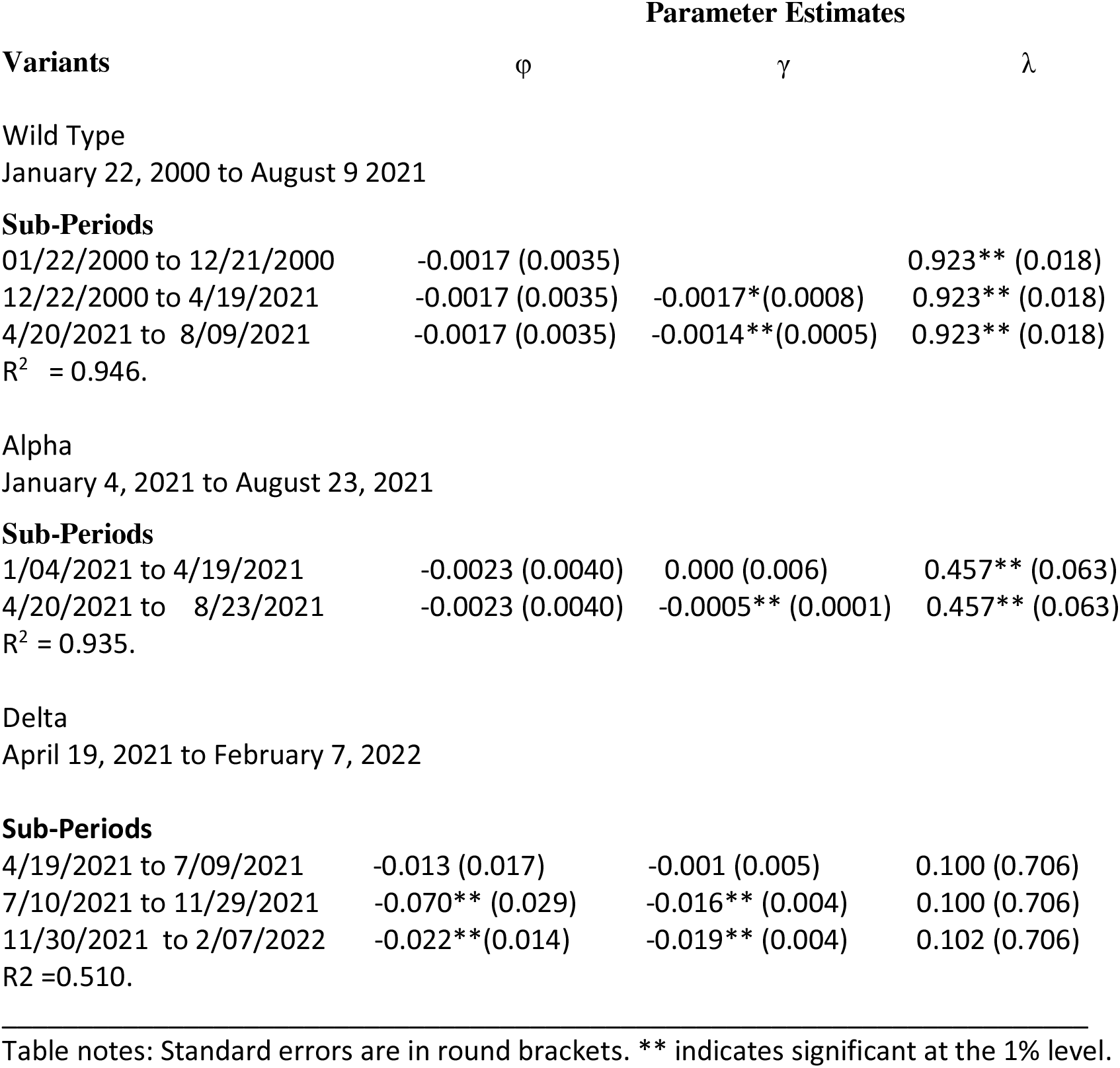
**Non-Linear IV Least Squares Parameter Estimates (Standard Error)** **For the Wild Type, Alpha and Delta Variants.**

| Variants | Parameter Estimates |  |  |
| --- | --- | --- | --- |
| | $\varphi$ | $\gamma$ | $\lambda$ |
| Wild Type<br>January 22, 2000 to August 9 2021 |  |  |  |
| **Sub-Periods** |  |  |  |
| 01/22/2000 to 12/21/2000 | -0.0017 (0.0035) |  | 0.923** (0.018) |
| 12/22/2000 to 4/19/2021 | -0.0017 (0.0035) | -0.0017*(0.0008) | 0.923** (0.018) |
| 4/20/2021 to 8/09/2021 | -0.0017 (0.0035) | -0.0014**(0.0005) | 0.923** (0.018) |
| $R^2 = 0.946$. | | | |
| Alpha<br>January 4, 2021 to August 23, 2021 |  |  |  |
| **Sub-Periods** |  |  |  |
| 1/04/2021 to 4/19/2021 | -0.0023 (0.0040) | 0.000 (0.006) | 0.457** (0.063) |
| 4/20/2021 to 8/23/2021 | -0.0023 (0.0040) | -0.0005** (0.0001) | 0.457** (0.063) |
| $R^2 = 0.935$. | | | |
| Delta<br>April 19, 2021 to February 7, 2022 |  |  |  |
| **Sub-Periods** |  |  |  |
| 4/19/2021 to 7/09/2021 | -0.013 (0.017) | -0.001 (0.005) | 0.100 (0.706) |
| 7/10/2021 to 11/29/2021 | -0.070** (0.029) | -0.016** (0.004) | 0.100 (0.706) |
| 11/30/2021 to 2/07/2022 | -0.022** (0.014) | -0.019** (0.004) | 0.102 (0.706) |
| $R^2 = 0.510$. | | | |

Table notes: Standard errors are in round brackets. \*\* indicates significant at the 1% level.

The results for the Alpha variant are similar. Previous cases have no significant impact on new cases but vaccines are effective only in the second sub-period. For both the Wild Type and Alpha model fit is very good with high R^2^ values. Unlike the other variants alpha never became dominant like Delta or Omicron which arrived later and eventually generated all the new cases. The number of alpha new cases reached a peak on May 3, 2021 at 16 per hundred thousand and then declined steadily after that. On that day the proportion of the population with one shot of the vaccine had reached 34% but only 3.3% of the population had ever been infected. It seems probable that the rapid deployment of the vaccine helped to arrest the growth of the alpha variant and contributed to its extinction or replacement by the Delta variety.

The results for Delta differ from the first two variants. The regression coefficients for previous infections, ϕ_δ,_ are significant for sub-periods 2 and 3. They are larger in absolute than for the other two variants and are larger in absolute value that the vaccine parameter estimates which have a significant negative effect on new cases as is the case for the other two variants.

There were other regressors used in the model. The Oxford Persistency Index is variable which attempts to measure the effects of various policies to combat Covid 19. It was seldom significant, perhaps because there are ten other actions included in the index so that the effects of masking and social distancing get diluted and appear not to be effective. Most Covid researchers believe that these two activities were important contributors to the decline of the virus.

Other variants were used as regressors and were sometimes significant but played a relatively small role in explaining new cases. New variants arrived at regular intervals and they displaced the existing variant so that a decline in new cases of the existing variant could be caused either by the effect of vaccines or because the variant was being replaced by a more infectious version of the virus. This process was quite rapid. For example, Delta was replaced completely by the omicron variant in about seven weeks. That variants are replaced so rapidly may seem strange but this behaviour is predicted by the simulation literature on SIR models with multiple infections^2^.

## Discussion

Some of the results presented above are somewhat unexpected. Because of that it is worth discussing what happens in the Alpha case in more detail. The regression coefficient for the vaccine variable is significant only in the second sub-period and is very small. There are two reasons for this. First, new cases per-capita are much smaller than vaccines per capita. However, the contribution of vaccines to the explanation of new cases is also very small. In Table 3 the relative contributions to the explanation of new Alpha cases are listed. This represented by the increases in both the ln-likelihood function and the R^2^ values.

**Table 3.** Decomposition of Explained Variation in Alpha New Cases.

| Variables Included | Ln(L) | $R^2$ |
| --- | --- | --- |
| Case 1 Baseline | -584.356 | 0.553 |
| Case 2 Baseline + $\text{Lag}(n_\alpha)$ | -377.639 | 0.981 |
| Case 3 Baseline + $\text{Lag}(n_\alpha)$<br>+ $X, S_w, S_\delta$ | -373.629 | 0.921 |
| Case 4 ALL | -348.822 | 0.935 |

Table Note. Base line variables are the cubic polynomial and the six day dummies

The baseline variables together with the lagged value of new Alpha cases explain most of the variation in new Alpha cases. While the model does a good job of explaining the data, as Figure 2 shows, the vaccine and previous cases explain just over one percent of the variation in new Alpha cases.

**Figure 2.**
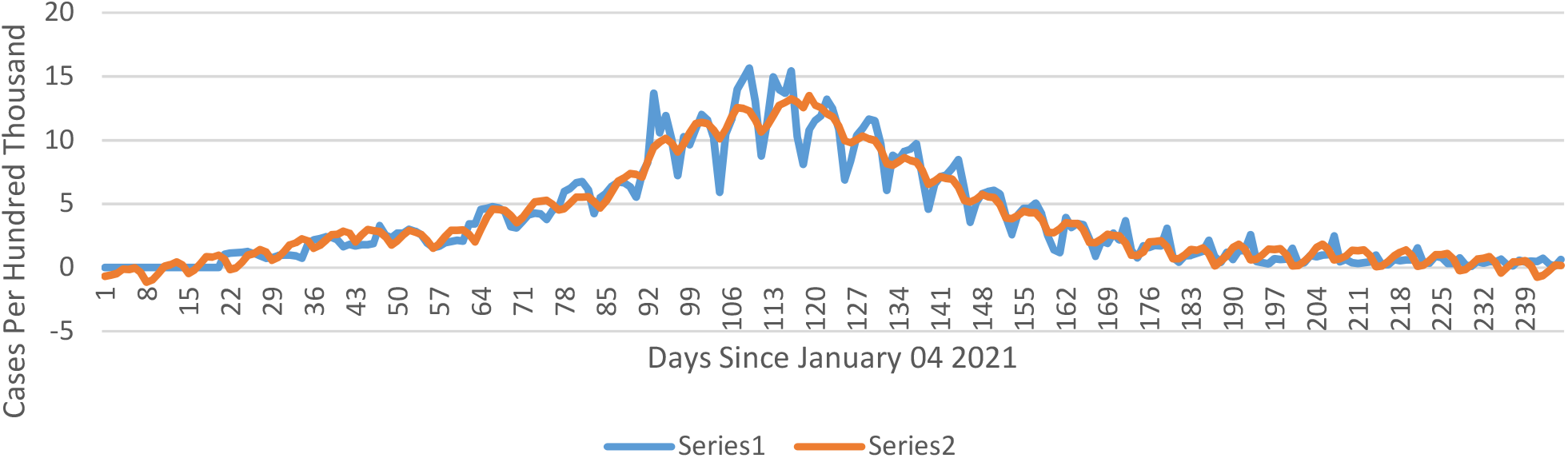
Series 1: Actual New Alpha Cases. Series 2: Predicted New Alpha Cases

ϕ_α_ is much larger in absolute value than γ_α_ for both sub-periods and is not significant; but there was no difference in the estimates of this parameter across the two sub-periods. This may be due to the fact that all the estimates of are IV estimates and these are consistent but biased in small samples and have much larger standard errors than least squares estimates. The consequence of this is that while the estimates γ_α_ are probably reliable the regression model described in equation (4) is not particularly informative about the impact of previous infections on new Alpha cases.

New cases for the Delta variant also display some surprises. First the γ_δ_ are very significant for the second and third sub-periods and are much larger smaller in absolute value than for the other two variants under consideration, an unexpected result since vaccines were reported to be more effective for Delta, in contrast to results reported in Sharma, Ketaki, Archana Koirala, Katrina Nicolopoulos et al (2021). ϕ_δ_ is negative, significant, and much larger in absolute value than any other of the vaccine coefficients. Previous Delta cases do help to prevent re-infection.

To see if previous cases of other variants had an effect on new cases *T*_*t*_ = ∑*T*_*vt*_ was used instead of *T*_*δ t*_ in equation (4). The regression coefficients associated with T_t_ were insignificant and positive or significantly positive, which is the wrong sign. This means that having been infected with other variants offers no immunity against the Delta variant and may actually increase the probability of contracting the Delta version of Covid. This experiment was carried out for the other two variants but the results were not conclusive. This is the main result of the paper and so far to the author’s knowledge has not been reported elsewhere. However, it should be interpreted cautiously for reason that are discussed below. It also means that the traditional^3^ view that heard immunity could be achieved if a large enough percentage of the population contracted the virus may not be correct. Variants are disappearing, not because the infected can’t find other individuals to infect but because they are being removed by the arrival of newer variants.

It is not clear what the implications of this result are. The serial nature of variant behaviour suggests that this might not matter very much. New variants eliminate existing variants completely so the fact that only previous cases of Delta, for example, provide protection against getting Delta a second time is not important if the replacement of Delta by Omicron means that the Delta variant never reappears. Delta was last seen in the United States on December 1, 2021 and so far (May 25, 2022) there have been no new cases of Delta. Only time will tell how this turns out.

As noted earlier, there is only partial protection against the Omicron variant from previous Omicron infections and no information on the prophylactic value of previous non-Omicron infections on Omicron. The CDC reports (May 22, 2022) that 57.9 % of new cases are of the BA2.12.1 variety and it is quickly being replaced by the variants, BA 4 and BA 5, which are even more contagious than earlier BA variants. Consequently, it is too early to determine the protective value of vaccines or previous infections making the consequences of evolution of the BA variants of Covid 19 in the United Stated very uncertain. It is also the case that the procedures used here will have difficulty in explaining the evolution of Omicron because of the low correlation between observed and actual new cases of the variant. Many new Omicron cases are generated by individuals using a self-administered antigen test whose results are never reported and there are many less serious case that are never even diagnosed.

Finally, two additional points are worth mentioning. Using aggregate data in equation (4) which does not take into account any information on individual variants does not fit the model so variants are clearly important and need to be analysed separately. On the other hand, the sequential analysis of individual variants also raises statistical problems. As is clear from Table 1, Delta started after the Wild Type ended so there is no variation in T_w_ in the three sub-periods for which the Delta variant was active. Thus it is impossible to make any inference about the effect of Wild Type infections on Delta new cases. Thus, as noted above, the result that previous other cases may not affect current new cases, a result that was determined by looking at the coefficient of T_t_ in equation (4) may be suspect. Because T_t_ has components with no variation this coefficient could be biased casting some doubt on the validity of the result. While the analysis of time series data provides useful information this is a major limitation for models which rely on the kind of data that is employed here.

## Data Availability

The data is publicly available on the Our World in Data website

## Footnotes

1 This was first reported by Lavezzo E., E. Franchin, C. Ciavarella et al. (2020).

2 See, for example, Zhao et al (2021)

3 The SIR model described by Hethcote (2001) is how many immunologists have modeled infectious diseases.

